# Impact of hyperactive neuropsychiatric symptoms on brain morphology in mild cognitive impairment and Alzheimer’s disease

**DOI:** 10.1101/2022.08.28.22279300

**Authors:** Lyna Mariam El Haffaf, Lucas Ronat, Adriana Cannizzaro, Alexandru Hanganu, for the ADNI

## Abstract

**Background:** The neuroanatomy of hyperactive neuropsychiatric symptoms (NPS) is poorly understood, and it is unclear whether these symptoms result from the same pathogenic processes responsible for cognitive decline or if they have an independent etiology to the neurodegeneration due to Alzheimer’s disease (AD).

**Objective:** We aim to investigate how the severity of hyperactive NPS (i.e., agitation, disinhibition, and irritability) can impact brain structures at different stages of cognitive decline.

**Methods:** Neuropsychiatric and 3T MRI data from 223 cognitively normal (CN) participants, 367 participants with mild cognitive impairment (MCI) and 175 participants with AD were extracted from the Alzheimer Disease Neuroimaging Initiative (ADNI) database. Statistical analyses based on the general linear model (GLM) were performed to define the effects of neuropsychiatric variables on brain structures in a two-by-two comparison (AD-MCI, CN-MCI and CN-AD). Linear regression analysis was also performed to investigate cortical changes as a function of NPS severity.

**Results:** In the AD group, the frontal dorsolateral is the most influenced region receiving an impact from more severe agitation, disinhibition and irritability. In AD, agitation and irritability influence some temporal inferior and parietal superior regions. Furthermore, severe disinhibition seems to have a stronger influence on CN participants compared to the other two groups, particularly in the occipital lingual, frontal middle rostral and frontal pars triangularis regions.

**Conclusion:** Our study shows that hyperactive NPS influence differently the brain morphology at different stages of cognitive performance. This might imply that their severity should be evaluated in relation to results of cognitive evaluations.

## INTRODUCTION

Alzheimer’s disease (AD) is a chronic neurodegenerative disease and the leading cause of dementia. As of 2016, 6.9% of Canadians aged 65 and older are living with diagnosed dementia [1]. The prevalence of this disease increases with age, and it affects more women than men [1]. In addition to the cognitive and functional impairment affecting these patients, neuropsychiatric symptoms (NPS) are often associated with the disease [2]. In fact, 97% of patients experience at least one NPS and around half of them experience four or more symptoms [3, 4]. NPS are a heterogeneous group of symptoms that contribute to cognitive and functional changes and can accelerate disease progression [4]. In fact, Peter et al. [5] showed that specific NPS including aggression, hallucination, delusions, depression, and anxiety are associated with shorter survival time from mild Alzheimer’s disease to severe dementia or death. Moreover, NPS are associated with a greater possibility of conversion to AD from mild cognitive impairment (MCI) [6, 7]. Thus, understanding these effects of NPS may be a lead for the prevention of the disease progression.

Drawing on the taxonomic concept commonly used in delirium [8], hyperactive symptoms of AD include agitation, disinhibition, and irritability. These symptoms represent one of the most difficult sets of symptoms to manage, causing an increase in institutionalization and a burden for caregivers [9]. The current study focuses on the hyperactive NPS (i.e., agitation, disinhibition, and irritability) for a few reasons. First, hyperactive symptoms can be grouped based on their onset. In fact, most hyperactive NPS appear at the MCI or pre-clinical stage of the disease, except for disinhibition, which has a later onset [10]. Second, although clustering studies have remained inconsistent in grouping co-occurring NPS, a few models clustered agitation, disinhibition, and irritability under the hyperactive disturbance cluster [11, 12]. Using confirmatory factor analysis on patients with AD, Cheng et al. [13] proposed a similar four factor clustering model which include behavioral problems (agitation, disinhibition, irritability and aberrant motor behavior), mood disturbance (apathy, depression, anxiety, sleep and appetite), psychosis (delusions and hallucinations) and euphoria. In addition, cluster studies have suggested that there is a likelihood of common underlying molecular and cellular pathologies for symptoms of the same cluster [9]. Thus, studies seem to consistently group hyperactive symptoms. Third, in addition to clustering studies, common brain regions are involved with hyperactive NPS in participants with AD. In fact, these symptoms exhibit a common deficit in the appropriate inhibition of action [9]. Along with these deficits, these symptoms seem to be related to increased functional connectivity of the anterior cingulate cortex (ACC) and the right insula of the salience network which leads to inappropriate behaviors [14, 15]. In addition, studies indicate that frontal brain regions and ACC interact with the ventral and dorsal striatal nuclei which mediate inhibition of impulsive thoughts and motor responses [16-18]. In line with these findings, MRI studies in AD show that agitation is related to greater atrophy of the frontal, cingulate, insular, amygdala and hippocampal region [19-21]. In addition, Hsu and colleagues [22] found that the severity of the agitation is correlated to the atrophy score of the posterior temporal lobe. Regarding disinhibition, studies reported its association with the ACC as well as the insula, the middle frontal lobe and bilateral cingulate cortex [6, 14]. Finally, irritability is associated with decreased volume of bilateral anterior insula and right posterior insula [23]. In sum, hyperactive symptoms are characterized by dysfunction of the orbitofrontal-subcortical circuit which connects frontal monitoring systems to the limbic system [24]. This suggests that there might be a common symptomatology among these three symptoms.

Thus, the aim of this study is to investigate how the severity of hyperactive symptoms can impact the cortical morphology at different stages of cognitive decline. Specifically, we will evaluate the changes in cortical volume as a primary objective, as well as cortical area and thickness, since ultimately, they determine the volume and are influenced by different factors [25, 26]. Previous studies also reported that cortical volume, thickness and area are key features in the diagnosis of MCI and AD [27]. We hypothesize that hyperactive symptoms share common cortical structures and the involved brain structures will be the same between cognitively normal (CN) individuals, participants with MCI and with AD.

## MATERIALS AND METHODS

### PARTICIPANTS

Participants were extracted from the Alzheimer’s Disease Neuroimaging Initiative (ADNI) database (adni.loni.usc.edu). The ADNI was launched in 2003 as a public-private partnership, led by Principal Investigator Michael W. Weiner, MD. The primary goal of ADNI has been to test whether serial magnetic resonance imaging (MRI), positron emission tomography, other biological markers, and clinical and neuropsychological assessment can be combined to measure the progression of MCI and early AD.

We extracted the structural MRI data of 765 participants, specifically: Cognitively Normal (CN) = 223, MCI = 367, AD = 175. All patients with AD met National Institute of Neurological and Communication Disorders/Alzheimer’s Disease and Related Disorders Association criteria for probable AD: (1) a Mini-Mental State Examination (MMSE) score between 20 and 26, (2) a global Clinical Dementia Rating of 0.5 or 1, (3) a sum-of-boxes Clinical Dementia Rating of 1.0 to 9.0. Therefore, all AD participants are only mildly impaired. Entry criteria for patients with amnestic MCI include: (1) a Mini-Mental State Examination score of 24 to 30 and (2) a Memory Box score of at least 0.5.

Python 3.9 with Pandas 1.1.2 library was used to compile data of participants. Excluding criteria were: (i) incomplete assessments, (ii) incomplete neuropsychiatric and neuropsychological assessments, (iii) presence of psychiatric history (major depression, schizophrenia, bipolar disorder, substance abuse, post-traumatic stress, obsessive-compulsive disorder), (iv) presence of neurological history (stroke, head injury, brain tumor, anoxia, epilepsy, alcohol dependence and Korsakoff, neurodevelopmental disorder), (v) prematurity, and (vi) diagnostic criteria in favor of other neurodegenerative or neurological etiology (Parkinson’s disease, frontotemporal degeneration, progressive supranuclear paralysis, corticobasal degeneration, Lewy body dementia, amyotrophic lateral sclerosis, multiple sclerosis, multi-system atrophy, vascular dementia). All data are available on the ADNI websites upon demand (http://adni.loni.usc.edu/data-samples/access-data/). Ethics committee approval and individual patient consents were received by the corresponding registration sites according to ADNI rules (http://adni.loni.usc.edu/methods/documents/). This study was approved by the Comité d’éthique de la recherche vieillissement-neuroimagerie CER VN 19-20-06.

### NEUROPSYCHIATRIC ASSESSMENT

Each participant underwent a neuropsychiatric assessment via the Neuropsychiatric Inventory (NPI). The NPI is a clinical questionnaire that assesses 12 behavioral and psychological symptoms of dementia (i.e., delusions, hallucinations, agitation, depression, anxiety, euphoria, apathy, disinhibition, irritability, aberrant motor behavior, nighttime behavior disorders and appetite disorders) [28]. The presence or absence and severity of each NPS is rated by the patient’s primary caregiver. The severity is rated on a 3-point scale where 1 is mild, 2 is moderate and 3 is marked source of behavioral disruption. Based on our hypothesis, we extracted the NPS required for our study: agitation/aggressiveness, disinhibition, and irritability. The values of severity of NPS were included in our statistical model.

### PROCEDURE

MRI data were processed on the Cedar cluster of the Digital Research Alliance of Canada (http://www.alliancecan.ca), on the CentOS Linux version 7, with FreeSurfer 7.1.1 (http://surfer.nmr.mgh.harvard.edu), Centos 7.0_x86_64 version, [29, 30] and automatically managed and verified by an in-house pipeline (github.com/alexhanganu/nimb). The results enabled the obtaining of volume, thickness and area of cortical regions of interest (ROI) based on the Desikan atlas [31]. Cortical thickness was computed as the average of (1) the distance from each white surface vertex to their corresponding closest point on the pial surface (not necessarily at a pial vertex) and (2) the distance from the corresponding pial vertex to the closest point on the white surface [32]. Cortical surface area was computed based on the triangular face of the surface representation with corresponding vertex coordinates *abc* of the corresponding triangle corner and dividing by two the vector norm of the cross product x of the differences between vertex coordinates: |*(a-c)* x *(b-c)*| */2* [33]. Cortical volume was based on defining oblique truncated triangular pyramids which were divided into three irregular tetrahedra and their volumes were calculated [34]. Each voxel in the normalized brain volume was assigned to one of 40 labels, using a probabilistic atlas obtained from a manually labeled training set [35]. Brain volumes were corrected using the Estimated Total Intracranial volume which is a metric computed from the amount of scaling based on the MNI305 space talairach transformation [36]. We used the regression-based correctional method, which was shown to provide advantages over the proportion method [37].

### STATISTICAL ANALYSIS

For this study, descriptive analyses were performed using SPSS version 26 software. These analyses verified the similarity of the groups in terms of gender distribution, age, years of education, the mean MMSE score and NPS severity (respectively mean comparisons by Student test and contingency Chi^2^ analysis). A whole-brain analysis was performed using general linear model (GLM) analysis with FreeSurfer mri_glmfit for each NPS of interest to assess their effect on brain’s cortical volume, thickness and area, between the groups (AD vs MCI, CN vs MCI, CN vs AD). Results underwent a Monte-Carlo correction with a vertex-level threshold of p<0.05. The severity of NPI subscale score was included as fixed factors, while MRI parameters (cortical volume, thickness and areas) were regarded as dependent variables. The Max-value provided by the GLM analysis indicates the difference in slope of cortical volume, surface area or thickness relative to NPS severity between two groups. Based on the results of the GLM analysis, ROI (as per Desikan atlas) underwent an additional linear regression analysis for intergroup comparison to visualize the cortical change as a function of neuropsychiatric symptom severity.

## RESULTS

Descriptive analyses showed that groups were similar according to age. Regarding the gender, there were less females than males in the MCI group compared to the two other groups. The AD group had a lower average number of years of education and a lower MMSE scores compared to both MCI and CN groups (table 1). The prevalence analysis of each NPS severity in each group, showed that irritability was the most prevalent among CN, MCI and AD participants for all severity groups (6.7%, 26.4% and 35.4% respectively). (Table 1).

**Table 1.** Demographic characteristics and prevalence of the severity according to groups.

| **Variables** |  | **CN**<br>**(N = 223)** | **MCI**<br>**(N = 367)** | **AD**<br>**(N= 175)** | **Chi-**<br>**square/F** | **p** |
| --- | --- | --- | --- | --- | --- | --- |
| **Female, n (%)** |  | 108 (48.4%) | 132 (36%) | 85 (48.6%) | 12.2 | <.01 |
| **Male, n (%)** |  | 115 (51.6%) | 235 (64%) | 90 (51.4%) |  |  |
| **Age (years)** |  | 75.8 ± 5.08 | 74.8 ± 7.34 | 75.5 ± 7.47 | 1.6 | 0.21 |
| **Education (years)** |  | 16.03 ± 2.85 | 15.65 ± 2.95 | 14.71 ± 3.08 | 10.3 | <.001 |
| **Mean MMSE Score** |  | 29.09 ± 1.00 | 27.09 ± 1.77 | 23.45 ± 2.03 | 578.8 | <.001 |
| **Agitation, n (%)** | 1 | 5 (2.24%) | 44 (12%) | 28 (16%) | 19.8 | <.001 |
|  | 2 | 1 (0.45%) | 20 (5.45%) | 12 (6.86%) |  |  |
|  | 3 | 0 (0%) | 2 (0.54%) | 3 (1.71%) |  |  |
| **Disinhibition, n (%)** | 1 | 1 (0.45%) | 20 (5.45%) | 20 (11.4%) | 16.3 | <.001 |
|  | 2 | 0 (0%) | 3 (0.82%) | 8 (4.57%) |  |  |
|  | 3 | 0 (0%) | 2 (0.54%) | 0 (0%) |  |  |
| **Irritability, n (%)** | 1 | 13 (5.83%) | 61 (16.6%) | 41 (23.4%) | 25.8 | <.001 |
|  | 2 | 2 (0.90%) | 34 (9.26%) | 18 (10.3%) |  |  |
|  | 3 | 0 (0%) | 2 (0.54%) | 3 (1.7%) |  |  |

**Legend:** CN = Cognitively normal; MCI = mild cognitive impairment, AD = Alzheimer's
disease; p = p-value; 1, 2, 3 values for Agitation, Disinhibition, Irritability = severity level

### INTER-GROUP GLM ANALYSIS

GLM based inter-group analysis revealed significant impact on brain’s morphology in the AD-MCI contrast for the three NPS studied (table 2).

**Table 2.**
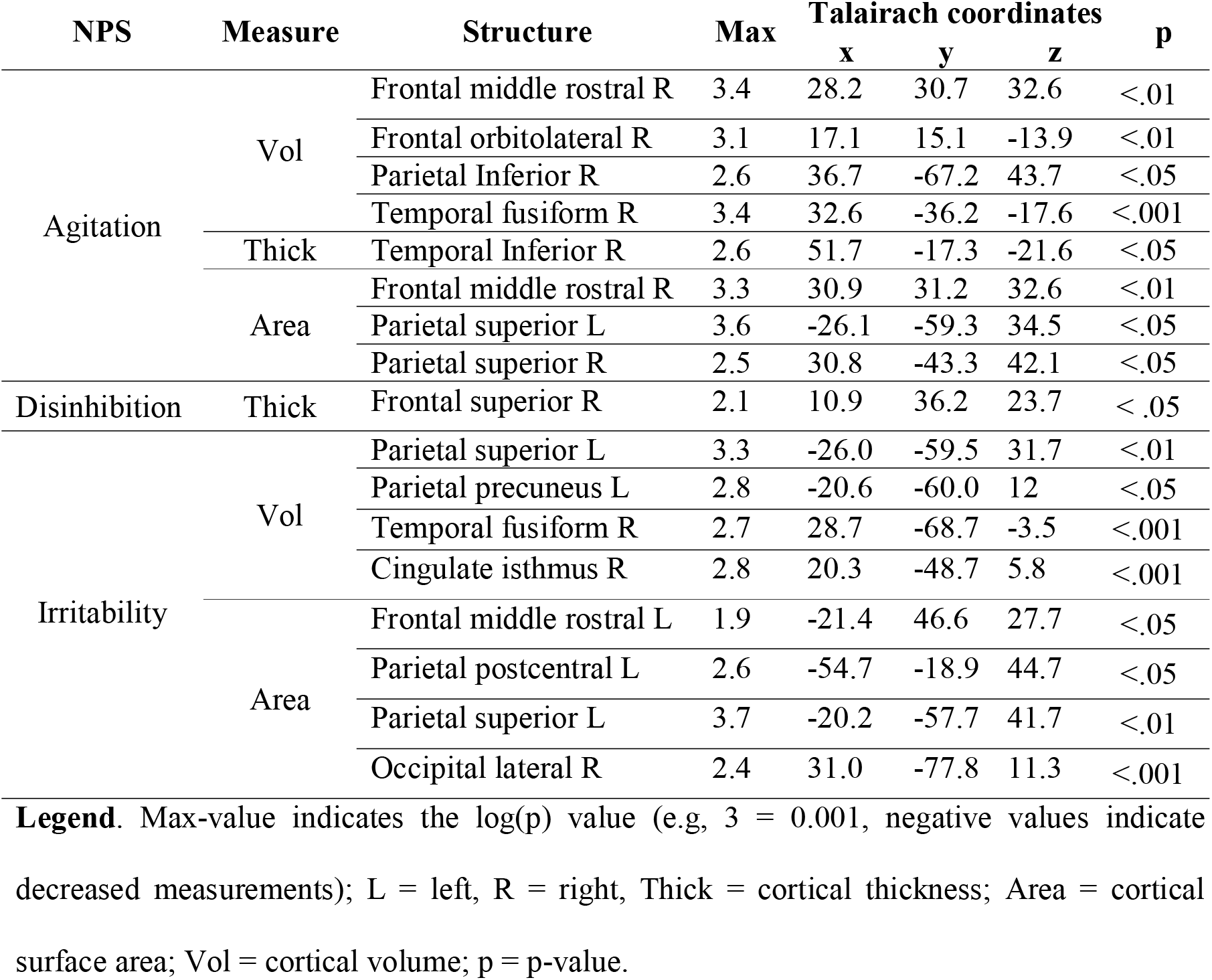
Inter-group (AD-MCI) analysis using the General linear model for the three NPS studied.

| NPS | Measure | Structure | Max | Talairach coordinates |  |  | p |
| --- | --- | --- | --- | --- | --- | --- | --- |
|  |  |  |  | x | y | z |  |
| Agitation | Vol | Frontal middle rostral R | 3.4 | 28.2 | 30.7 | 32.6 | <.01 |
|  |  | Frontal orbitolateral R | 3.1 | 17.1 | 15.1 | -13.9 | <.01 |
|  |  | Parietal Inferior R | 2.6 | 36.7 | -67.2 | 43.7 | <.05 |
|  |  | Temporal fusiform R | 3.4 | 32.6 | -36.2 | -17.6 | <.001 |
|  | Thick | Temporal Inferior R | 2.6 | 51.7 | -17.3 | -21.6 | <.05 |
|  | Area | Frontal middle rostral R | 3.3 | 30.9 | 31.2 | 32.6 | <.01 |
|  |  | Parietal superior L | 3.6 | -26.1 | -59.3 | 34.5 | <.05 |
|  |  | Parietal superior R | 2.5 | 30.8 | -43.3 | 42.1 | <.05 |
| Disinhibition | Thick | Frontal superior R | 2.1 | 10.9 | 36.2 | 23.7 | < .05 |
| Irritability | Vol | Parietal superior L | 3.3 | -26.0 | -59.5 | 31.7 | <.01 |
|  |  | Parietal precuneus L | 2.8 | -20.6 | -60.0 | 12 | <.05 |
|  |  | Temporal fusiform R | 2.7 | 28.7 | -68.7 | -3.5 | <.001 |
|  |  | Cingulate isthmus R | 2.8 | 20.3 | -48.7 | 5.8 | <.001 |
|  | Area | Frontal middle rostral L | 1.9 | -21.4 | 46.6 | 27.7 | <.05 |
|  |  | Parietal postcentral L | 2.6 | -54.7 | -18.9 | 44.7 | <.05 |
|  |  | Parietal superior L | 3.7 | -20.2 | -57.7 | 41.7 | <.01 |
|  |  | Occipital lateral R | 2.4 | 31.0 | -77.8 | 11.3 | <.001 |

**Legend.** Max-value indicates the log(p) value (e.g, 3 = 0.001, negative values indicate decreased measurements); L = left, R = right, Thick = cortical thickness; Area = cortical surface area; Vol = cortical volume; p = p-value.

Specifically, the frontal lobe, the superior, middle rostral and orbital regions were influenced by an increased severity of agitation, disinhibition and irritability. Agitation had an impact on the volume and surface area, disinhibition impacted the cortical thickness while irritability changed only the surface area. In the temporal lobe, significant impact was also depicted: AD participants have a stronger association between agitation severity and the volume and thickness of the right temporal inferior and fusiform region, as well as between irritability severity and the volume of the right temporal fusiform region. Finally, changes were also found in the parietal lobe in relation to agitation and irritability. In fact, a greater association was found between agitation severity and the surface area of the right inferior region, between irritability severity and the surface area of the postcentral region and between both agitation and irritability and the volumes of the bilateral superior region. Also, a stronger association was found between irritability severity and the volumes of the precuneus and cingulate isthmus regions. The occipital lobe was only impacted with respect to the surface area in the lateral region due to irritability.

In the CN-MCI comparison, only disinhibition showed significant results (table 3). Specifically, it was strongly associated to the volumes of the left frontal middle rostral, surface area of the right frontal pars triangularis and volumes of bilateral temporal inferior and right occipital lingual regions.

**Table 3.**
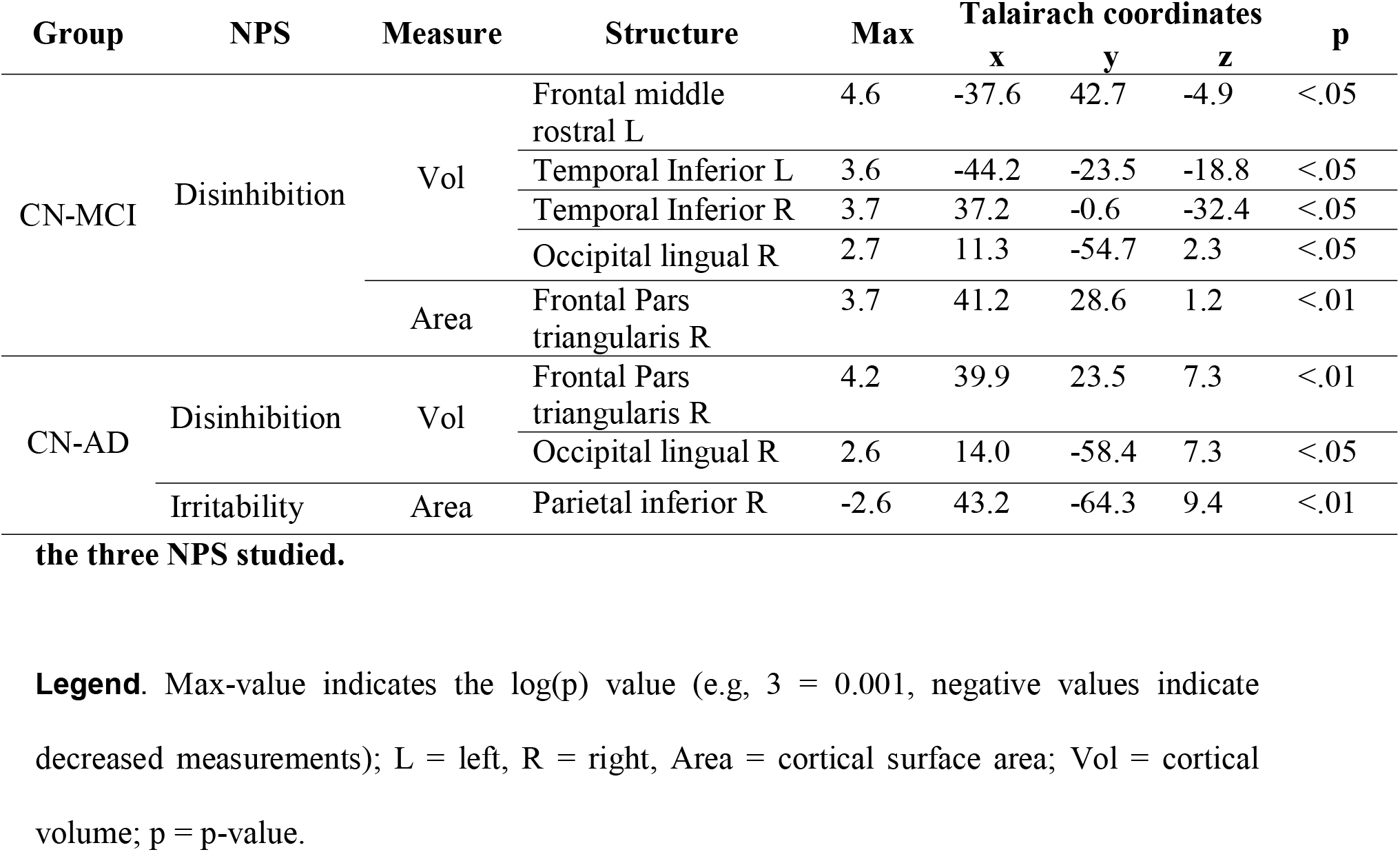
Inter-group (CN-MCI and CN-AD) analysis using the General linear model for the three NPS studied.

| Group | NPS | Measure | Structure | Max | Talairach coordinates |  |  | p |
| --- | --- | --- | --- | --- | --- | --- | --- | --- |
|  |  |  |  |  | x | y | z |  |
| CN-MCI | Disinhibition | Vol | Frontal middle rostral L | 4.6 | -37.6 | 42.7 | -4.9 | <.05 |
|  |  |  | Temporal Inferior L | 3.6 | -44.2 | -23.5 | -18.8 | <.05 |
|  |  |  | Temporal Inferior R | 3.7 | 37.2 | -0.6 | -32.4 | <.05 |
|  |  |  | Occipital lingual R | 2.7 | 11.3 | -54.7 | 2.3 | <.05 |
|  |  | Area | Frontal Pars triangularis R | 3.7 | 41.2 | 28.6 | 1.2 | <.01 |
| CN-AD | Disinhibition | Vol | Frontal Pars triangularis R | 4.2 | 39.9 | 23.5 | 7.3 | <.01 |
|  |  |  | Occipital lingual R | 2.6 | 14.0 | -58.4 | 7.3 | <.05 |
|  | Irritability | Area | Parietal inferior R | -2.6 | 43.2 | -64.3 | 9.4 | <.01 |

Interestingly, for the CN-AD inter-group comparison, disinhibition and irritability showed significant results, while agitation did not (table 3, Figure 1). In fact, disinhibition influenced positively the volumes of the right frontal pars triangularis while irritability showed a negative association with the surface area of the right parietal inferior region, implying in this case a greater relationship for AD and not for CN. Furthermore, disinhibition influenced strongly the right occipital lingual region in this contrast which is the same region that was depicted in the CN-MCI contrast for this NPS.

**Fig 1.**
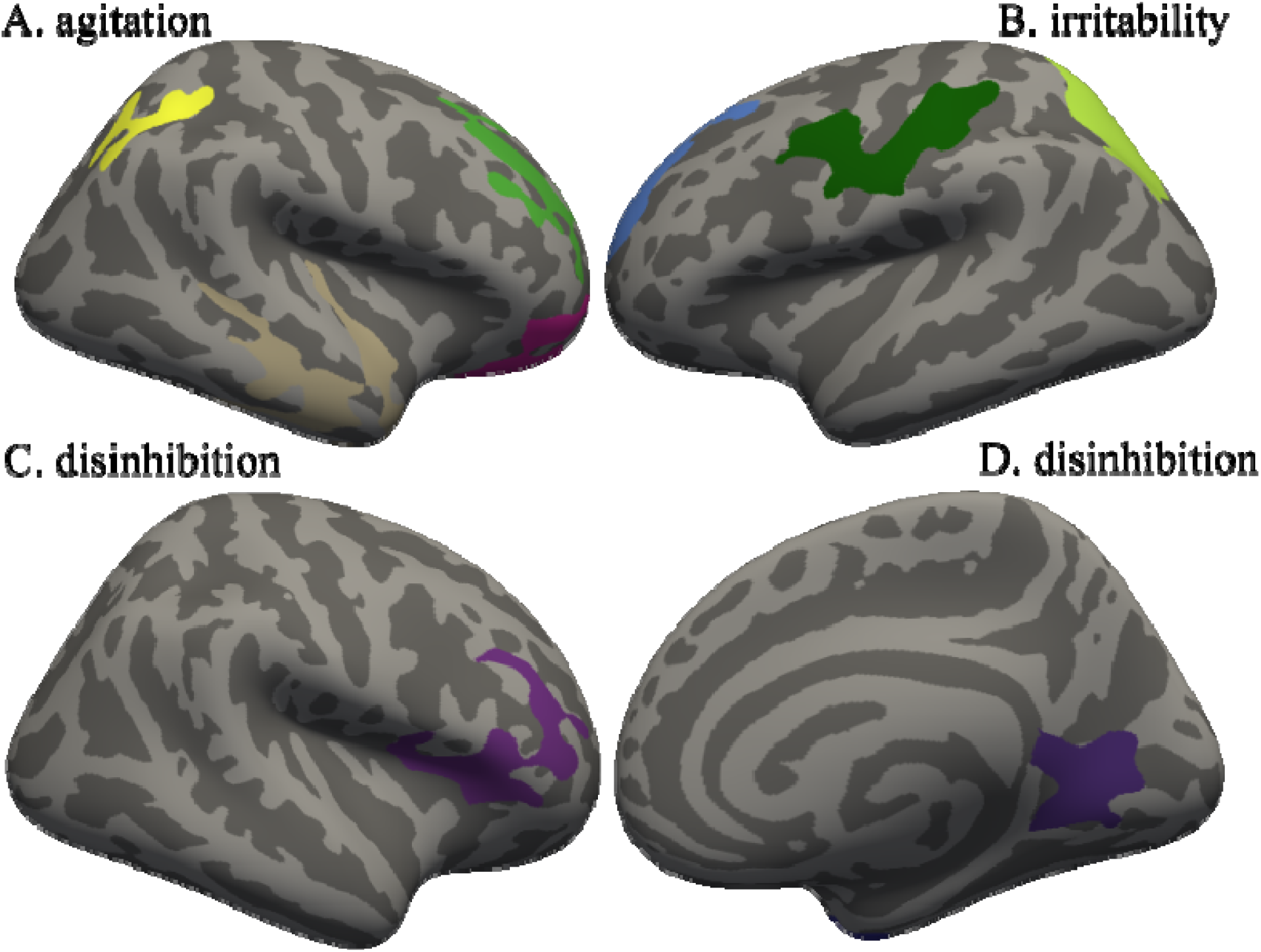
Regions of significant correlations for the inter-group analysis. **Legend:** Agitation (A) and the volume of the right temporal fusiform (grey), frontal middle rostral (green), frontal orbitolateral (pink) and parietal inferior (yellow) regions (AD-MCI); Irritability (B) and the area of the left frontal middle rostral (bleu), parietal superior (yellow) and parietal postcentral (green) regions (AD-MCI); Disinhibition (C) and the volume of the right frontal pars triangularis (purple) (CN-AD); Disinhibition (D) and the volume of the right occipital lingual region (purple) (CN-MCI).

### INTER-GROUP LINEAR REGRESSION ANALYSIS

The linear regression analysis allowed us to visualize the relationship between the cortical structures and the severity of these NPS. Only the comparison between the AD-MCI inter-group was retained because of the low prevalence of presence of the NPS studied in CN participants. Our results revealed a greater increase in the ROI for AD participants compared to MCI with higher severity scores for the three hyperactive symptoms (Fig. 2).

**Fig 2.**
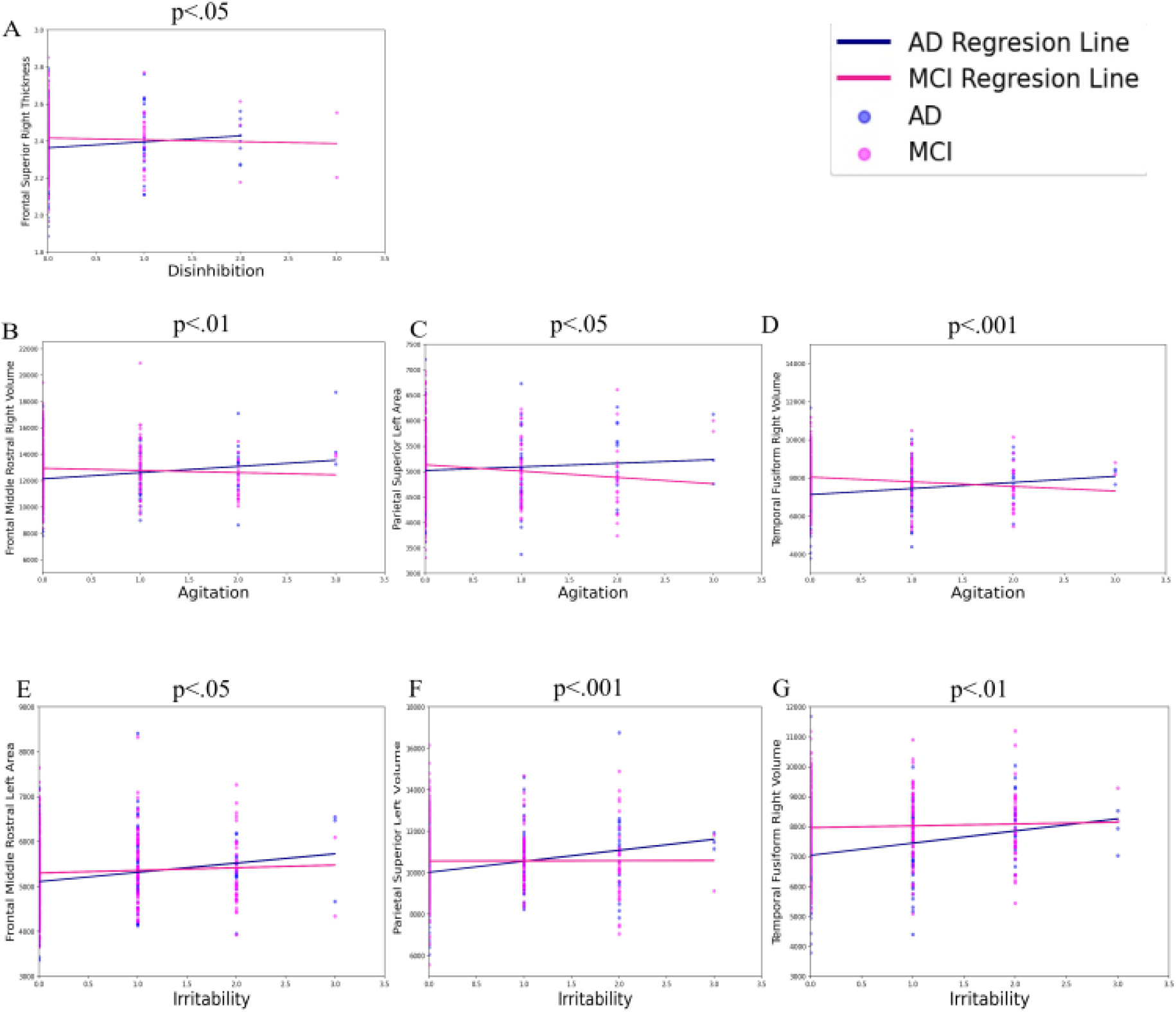
Intergroup linear regressions for AD-MCI inter-group comparison. **Legend:** MCI in pink; AD in blue; Cortical thickness of the (A) right frontal superior region as a function of disinhibition severity; Volume of the (B) right frontal middle rostral, surface area of the (C) left parietal superior and volume of the (D) right temporal fusiform regions as a function of agitation severity; Surface area of the (E) left frontal middle rostral, volume of the (F) left parietal superior and (G) right temporal fusiform regions as a function of irritability severity.

## DISCUSSION

The aim of this study was to examine the impact of NPS from the hyperactive symptoms cluster, on brain morphology, at different stages of cognitive performance. We showed that, in the AD group, severe agitation seems to impact mainly the frontal and temporal regions, disinhibition influences only the frontal superior regions, while irritability has a greater impact on the brain, inducing changes in almost all lobes. Functionally, in AD, the most impact is present in the frontal dorsolateral regions (superior and middle rostral regions), shared amongst the three hyperactive symptoms while the temporal inferior (fusiform and inferior regions) and the parietal superior regions are shared amongst agitation and irritability. Furthermore, disinhibition seems to have a stronger influence in the CN participants in comparison to the other two groups. Specifically, it showed a greater association in the occipital lingual region in both the CN-MCI and the CN-AD contrasts, and an impact on the frontal middle rostral and frontal pars triangularis in the CN-MCI and CNAD contrasts.

First, AD-MCI inter-group comparison revealed that the frontal dorsolateral regions (superior and middle rostral regions) are shared amongst the three hyperactive symptoms while the temporal inferior (fusiform and inferior regions) and the parietal superior regions are shared amongst agitation and irritability NPS. These results are consistent with findings from prior studies which highlight the role of orbitofrontal-subcortical circuits in personality changes including agitation, disinhibition and irritability [14, 19, 20, 24, 38]. Specifically, research show an association between agitation and the frontal region, the anterior and posterior cingulate cortex, the insula, amygdala and hippocampus [21]. We also found that the volume of the right temporal inferior region was associated with agitation. This can be supported by Hsu et al. [22] who found a similar association between agitation and the right temporal posterior lobe. In frontotemporal dementia, the right sided frontotemporal degeneration is associated with socially undesirable behavior which provides further support for the implication of this cortical structure in hyperactive symptoms at the AD stage [39]. Regarding disinhibition, we found an association between this symptom and the cortical thickness of the right frontal superior region. Similarly, studies found disinhibition to be associated with the right frontal middle gyrus and the ACC which are regions thought to be implicated in the regulation of behavior [6, 40]. Moreover, patients with frontotemporal dementia often exhibit disinhibition [41]. Altogether, these studies suggest the implication of the frontal cortex in disinhibition. Another result of interest that emerged from our analysis was that agitation and disinhibition were mostly associated with changes in the right hemisphere. While the relation between regulation of behavior and the right hemisphere is not completely established, our results are in line with studies showing the role of the right hemisphere in the regulation of social and emotional behavior [40]. Finally, irritability was associated with frontal, parietal, temporal inferior and cingulate changes. Moon et al. [23] found that irritability was associated with anterior and posterior insula changes. Also, using diffusion tensor imaging, Tighe et al. [15] found that irritability was associated with decrease white matter integrity in the anterior cingulate cortex. Moreover, in preclinical Huntington disease patients and children with irritability, the parietal lobe may be involved in attention tasks to maintain emotional and/or behavioral response in frustrating condition [42, 43]. There seem to be a link between irritability and the insula and the cingulate cortex which is in part observed in this study. Thus, in this intergroup comparison, frontal regions which are implicated with the regulation of behavior are found to be associated with the three NPS studied.

For CN-MCI comparison, only disinhibition revealed significant results, showing implication of the frontal and temporal regions with this NPS. Finally, for the CN-AD intergroup comparison, significant results were found for disinhibition and irritability but not agitation. Interestingly, the volume of the right occipital lingual and the volume and surface area of the right frontal pars triangularis regions was found in both CN-MCI and CN-AD contrast for disinhibition. Confirming our hypothesis, results suggests that hyperactive symptoms share common cortical structures. Specifically, we observed an association between hyperactive NPS and the frontal cortex. This is consistent with previous studies which outline the role of the orbitofrontal-subcortical circuit in hyperactive symptoms. However, for the same NPS, brain structures are not the same when comparing different group contrast (CN-AD to CN-MCI and to AD-MCI). Overall, these results suggest that, at early stages of AD, frontal cortices are already involved with hyperactive NPS.

Additional linear regression analysis was done to visualize GLM results. Our results revealed a greater increase in all ROI for AD participants compared to MCI with higher severity scores for the three hyperactive symptoms. This is a surprising finding, considering that most studies find that with increasing severity, there is a decrease in ROI. A possible explanation for this is chronic neuroinflammation associated with activated microglia cells and an increase of serum proinflammatory cytokines which contribute to an increase rate of cognitive decline in AD [44-46]. In particular, agitation is associated with increased pro-inflammatory cytokines (systemic tumor necrosis factor alpha (TNFα)) detected in the serum of AD patients [47]. Also, higher levels of anti-inflammatory cytokine IL-10 in the cerebrospinal fluid (CSF) has been associated with lower agitation NPI scores in patients with dementia [48]. Holmgren et al. [48] suggested that this finding indicates a protective effect on mental health by a strong anti-inflammatory response that can balance inflammation in the AD brain. To our knowledge, no studies found significant correlations between disinhibition and irritability and inflammation markers. However, since there seems to be brain regions shared between hyperactive NPS, it might be possible that the increase in ROI observed is due to the same neuroinflammatory processes involved in agitation. Another explanation to these results is the hypothesis that hyperactive symptoms in AD may result from compensatory overactivity of the noradrenergic system, secondary to locus coeruleus neuronal loss [49]. This compensatory central overactivity could shift noradrenergic actions in the frontal cortices resulting in impaired attention, disinhibition, and affective dysregulation [50]. Finally, research show that AD pathology tends to progress from the ventral to dorsal cortical areas indicating a later spread of pathology to frontal regions [9]. Since the ADNI database included more participants with earlier stages of AD (MCI and mild AD), cortical regions associated with hyperactive symptoms may be compensating neuronal loss in other regions of the brain. Lastly, there seems to be more marked damage in more severe stages of NPS studied which is shown by the steeper linear regression in AD-MCI group comparison.

### STRENGTHS AND LIMITATIONS

This study sheds light on the involvement of hyperactive symptoms on neuroanatomical correlates in AD. To our knowledge, this is the first time that intergroup comparison analyses have been performed to investigate the impact of hyperactive symptom severity on brain morphology. The ADNI database provided a large sample size and included all diagnostic groups across the AD spectrum allowing for intergroup analyses. These analyses led to the finding that these three NPS share common brain regions and that with an increase in their severity, there is an increase in brain volume, thickness, or area. Furthermore, this is one of the few studies to investigate the role of hyperactive NPS in CN participants. Altogether, this helps to draw a better picture of the impact of hyperactive symptoms at different stages of AD.

A few limitations to our study need to be addressed. Specifically, the prevalence of the NPS in the CN group was low (<6%), which influences the results referring to this group. Also, our study is cross-sectional and thus, no causal relationship can be concluded from the significant correlation we found. In fact, results observed could be due to the pathophysiological decline of AD which is correlated to symptom severity and thus, the more advanced the decline is, the more likely NPS are to be severe. Conversely, it is possible that with increasing NPS severity, the decline is greater, and therefore, the probability of belonging to the AD group rather than the MCI group is also greater. A longitudinal study would be needed to deepen the understanding of these results.

### CONCLUSION & IMPLICATION OF THE CURRENT STUDY

To our knowledge, this study is one of the few to focus on the cognitively normal stage and it also sheds light on the involvement of hyperactive symptoms on neuroanatomical correlates in AD. As hypothesized, we found some similar brain regions involved in hyperactive symptoms. We also found an increase in cortical volume, thickness or area associated with hyperactive symptoms. Furthermore, previous studies reported that cortical thickness and area are key features in the diagnosis of MCI and AD [27], while we also show that volumetry is an additional parameter that appears to show changes without corresponding significant changes in other parameters. Our study shows that hyperactive NPS influence differently the brain morphology at different stages of cognitive performance. This might imply that their severity should be evaluated in relation to results of cognitive evaluations.

## Data Availability

All data produced in the present work are contained in the manuscript.

## Abbreviations

ADNI: Alzheimer’s Disease Neuroimaging Initiative;
AD: Alzheimer’s Disease;
CN: Cognitively Normal;
MCI: Mild Cognitive Impairment;
GLM: General Linear Model;
NPI: Neuropsychiatric Inventory;
NPS: Neuropsychiatric Symptoms.

## FUNDING

This work was supported by a research bursary from NiEmoLab to Lyna El Haffaf, a doctoral research scholarship *Centre de Recherche de l’Institut Universitaire de Gériatrie Montréal (CRIUGM)*-Volet B in collaboration with NiEmoLab and a Faculty of Medicine of the Université de Montréal merit scholarship in collaboration with NiEmoLab given to Lucas Ronat, as well as funding from the Parkinson Canada-Parkinson Quebec (2018-00355); IUGM Foundation; *Fonds de Recherche du Québec Santé*; Lemaire Foundation given to Alexandru Hanganu.

## COMPETING INTERESTS

The authors have no conflict of interest to report.

## USE OF ADNI DATA

Data collection and sharing for this project was funded by the Alzheimer’s Disease Neuroimaging Initiative (ADNI) (National Institutes of Health Grant U01 AG024904) and DOD ADNI (Department of Defense award number W81XWH-12-2-0012). ADNI is funded by the National Institute on Aging, the National Institute of Biomedical Imaging and Bioengineering, and through generous contributions from the following: AbbVie, Alzheimer’s Association; Alzheimer’s Drug Discovery Foundation; Araclon Biotech; BioClinica, Inc.; Biogen; Bristol-Myers Squibb Company; CereSpir, Inc.; Cogstate; Eisai Inc.; Elan Pharmaceuticals, Inc.; Eli Lilly and Company; EuroImmun; F. Hoffmann-La Roche Ltd and its affiliated company Genentech, Inc.; Fujirebio; GE Healthcare; IXICO Ltd.; Janssen Alzheimer Immunotherapy Research & Development, LLC.; Johnson & Johnson Pharmaceutical Research & Development LLC.; Lumosity; Lundbeck; Merck & Co., Inc.; Meso Scale Diagnostics, LLC.; NeuroRx Research; Neurotrack Technologies; Novartis Pharmaceuticals Corporation; Pfizer Inc.; Piramal Imaging; Servier; Takeda Pharmaceutical Company; and Transition Therapeutics. The Canadian Institutes of Health Research is providing funds to support ADNI clinical sites in Canada. Private sector contributions are facilitated by the Foundation for the National Institutes of Health (www.fnih.org). The grantee organization is the Northern California Institute for Research and Education, and the study is coordinated by the Alzheimer’s Therapeutic Research Institute at the University of Southern California. ADNI data are disseminated by the Laboratory for NeuroImaging at the University of Southern California.

## APPENDIX 1

### ADNI Co-investigator

Michael Weiner, MD (UC San Francisco, PI of ADNI), Paul Aisen, MD (UC San Diego, ADCS PI and Director of Coordinating Center Clinical Core), Michael Weiner, MD (UC San Francisco, Executive Committee), Paul Aisen, MD (UC San Diego, Executive Committee), Ronald Petersen, MD, PhD (Mayo Clinic, Rochester, Executive Committee), Clifford R. Jack, Jr., MD (Mayo Clinic, Rochester, Executive Committee), William Jagust, MD (UC Berkeley, Executive Committee), John Q. Trojanowki, MD, PhD (U Pennsylvania, Executive Committee), Arthur W. Toga, PhD (UCLA, Executive Committee), Laurel Beckett, PhD (UC Davis, Executive Committee), Robert C. Green, MD, MPH (Brigham and Women’s Hospital/Harvard Medical School, Executive Committee), Andrew J. Saykin, PsyD (Indiana University, Executive Committee), John Morris, MD (Washington University St. Louis, Executive Committee), Enchi Liu, PhD (Janssen Alzheimer Immunotherapy, ADNI 2 Private Partner Scientific Board (Chair), Robert C. Green, MD, MPH (Brigham and Women’s Hospital/Harvard Medical School, Data and Publication Committee (Chair)), Tom Montine, MD, PhD (University of Washington, Resource Allocation Review Committee), Ronald Petersen, MD, PhD (Mayo Clinic, Rochester, Clinical Core Leaders (Core PI)), Paul Aisen, MD (UC San Diego, Clinical Core Leaders), Anthony Gamst, PhD (UC San Diego, Clinical Informatics and Operations), Ronald G. Thomas, PhD (UC San Diego, Clinical Informatics and Operations), Michael Donohue, PhD (UC San Diego, Clinical Informatics and Operations), Sarah Walter, MSc (UC San Diego, Clinical Informatics and Operations), Devon Gessert (UC San Diego, Clinical Informatics and Operations), Tamie Sather (UC San Diego, Clinical Informatics and Operations), Laurel Beckett, PhD (UC Davis, Biostatistics Core Leaders and Key Personnel (Core PI)), Danielle Harvey, PhD (UC Davis, Biostatistics Core Leaders and Key Personnel), Anthony Gamst, PhD (UC San Diego, Biostatistics Core Leaders and Key Personnel), Michael Donohue, PhD (UC San Diego, Biostatistics Core Leaders and Key Personnel), John Kornak, PhD (UC Davis, Biostatistics Core Leaders and Key Personnel), Clifford R. Jack, Jr., MD (Mayo Clinic, Rochester, MRI Core Leaders and Key Personnel (Core PI)), Anders Dale, PhD (UC San Diego, MRI Core Leaders and Key Personnel), Matthew Bernstein, PhD (Mayo Clinic, Rochester, MRI Core Leaders and Key Personnel), Joel Felmlee, PhD (Mayo Clinic, Rochester, MRI Core Leaders and Key Personnel), Nick Fox, MD (University of London, MRI Core Leaders and Key Personnel), Paul Thompson, PhD (UCLA School of Medicine, MRI Core Leaders and Key Personnel), Norbert Schuff, PhD (UCSF MRI, MRI Core Leaders and Key Personnel), Gene Alexander, PhD (Banner Alzheimer’s Institute, MRI Core Leaders and Key Personnel), Charles DeCarli, MD (UC Davis, MRI Core Leaders and Key Personnel), William Jagust, MD (UC Berkeley, PET Core Leaders and Key Personnel (Core PI)), Dan Bandy, MS, CNMT (Banner Alzheimer’s Institute, PET Core Leaders and Key Personnel), Robert A. Koeppe, PhD (University of Michigan, PET Core Leaders and Key Personnel), Norm Foster, MD (University of Utah, PET Core Leaders and Key Personnel), Eric M. Reiman, MD (Banner Alzheimer’s Institute, PET Core Leaders and Key Personnel), Kewei Chen, PhD (Banner Alzheimer’s Institute, PET Core Leaders and Key Personnel), Chet Mathis, MD (University of Pittsburgh, PET Core Leaders and Key Personnel), John Morris, MD (Washington University St. Louis, Neuropathology Core Leaders), Nigel J. Cairns, PhD, (MRCPath Washington University St. Louis, Neuropathology Core Leaders), Lisa Taylor-Reinwald, BA, HTL (Washington University St. Louis, Neuropathology Core Leaders), J.Q. Trojanowki, MD, PhD (UPenn School of Medicine, Biomarkers Core Leaders and Key Personnel (Core PI)), Les Shaw, PhD (UPenn School of Medicine, Biomarkers Core Leaders and Key Personnel), Virginia M.Y. Lee, PhD, MBA (UPenn School of Medicine, Biomarkers Core Leaders and Key Personnel), Magdalena Korecka, PhD (UPenn School of Medicine, Biomarkers Core Leaders and Key Personnel), Arthur W. Toga, PhD (UCLA, Informatics Core Leaders and Key Personnel (Core PI)), Karen Crawford (UCLA, Informatics Core Leaders and Key Personnel), Scott Neu, PhD (UCLA, Informatics Core Leaders and Key Personnel), Andrew J. Saykin, PsyD (Indiana University, Genetics Core Leaders and Key Personnel), Tatiana M. Foroud, PhD (Indiana University, Genetics Core Leaders and Key Personnel), Steven Potkin, MD (UC Irvine, Genetics Core Leaders and Key Personnel), Li Shen, PhD (Indiana University, Genetics Core Leaders and Key Personnel), Zaven Kachaturian, PhD (Khachaturian, Radebaugh & Associates (KRA), Inc, Early Project Development), Richard Frank, MD, PhD (General Electric, Early Project Development), Peter J. Snyder, PhD (University of Connecticut, Early Project Development), Susan Molchan, PhD (National Institute on Aging/National Institutes of Health, Early Project Development), Jeffrey Kaye, MD (Oregon Health and Science University, Site Investigator), Joseph Quinn, MD (Oregon Health and Science University, Site Investigator), Betty Lind, BS (Oregon Health and Science University, Site Investigator), Sara Dolen, BS (Oregon Health and Science University, Past Site Investigator), Lon S. Schneider, MD (University of Southern California, Site Investigator), Sonia Pawluczyk, MD (University of Southern California, Site Investigator), Bryan M. Spann, DO, PhD (University of Southern California, Site Investigator), James Brewer, MD, PhD (University of California--San Diego, Site Investigator), Helen Vanderswag, RN (University of California--San Diego, Site Investigator), Judith L. Heidebrink, MD, MS, (University of Michigan, Site Investigator), Joanne L. Lord, LPN, BA, CCRC (University of Michigan, Site Investigator), Ronald Petersen, MD, PhD (Mayo Clinic, Rochester, Site Investigator), Kris Johnson, RN (Mayo Clinic, Rochester, Site Investigator), Rachelle S. Doody, MD, PhD (Baylor College of Medicine, Site Investigator), Javier Villanueva-Meyer, MD (Baylor College of Medicine, Site Investigator), Munir Chowdhury, MBBS, MS (Baylor College of Medicine, Site Investigator), Yaakov Stern, PhD (Columbia University Medical Center, Site Investigator), Lawrence S. Honig, MD, PhD (Columbia University Medical Center, Site Investigator), Karen L. Bell, MD, (Columbia University Medical Center, Site Investigator), John C. Morris, MD (Washington University, St. Louis, Site Investigator), Beau Ances, MD (Washington University, St. Louis, Site Investigator), Maria Carroll, RN, MSN (Washington University, St. Louis, Site Investigator), Sue Leon, RN, MSN (Washington University, St. Louis, Site Investigator), Mark A. Mintun, MD (Washington University, St. Louis, Past Site Investigator), Stacy Schneider, APRN, BC, GNP (Washington University, St. Louis, Past Site Investigator), Daniel Marson, JD, PhD (University of Alabama – Birmingham, Site Investigator), Randall Griffith, PhD, ABPP (University of Alabama – Birmingham, Site Investigator), David Clark, MD (University of Alabama – Birmingham, Site Investigator), Hillel Grossman, MD (Mount Sinai School of Medicine, Site Investigator), Effie Mitsis, PhD (Mount Sinai School of Medicine, Site Investigator), Aliza Romirowsky, BA (Mount Sinai School of Medicine, Site Investigator), Leyla deToledo-Morrell, PhD (Rush University Medical Center, Site Investigator), Raj C. Shah, MD (Rush University Medical Center, Site Investigator) (Wein Center, Site Investigator), Ranjan Duara, MD (Wein Center, Site Investigator), Daniel Varon, MD (Wein Center, Site Investigator), Peggy Roberts, CNA (Wein Center, Site Investigator), Marilyn Albert, PhD (Johns Hopkins University, Site Investigator), Chiadi Onyike, MD, MHS (Johns Hopkins University, Site Investigator), Stephanie Kielb MD (Johns Hopkins University, Site Investigator), Henry Rusinek, PhD (New York University, Site Investigator), Mony J de Leon, EdD (New York University, Site Investigator), Lidia Glodzik, MD, PhD (New York University, Site Investigator), Susan De Santi, PhD (New York University, Past Site Investigator), P. Murali Doraiswamy, MD (Duke University Medical Center, Site Investigator), Jeffrey R. Petrella, MD (Duke University Medical Center, Site Investigator), R. Edward Coleman, MD (Duke University Medical Center, Site Investigator), Steven E. Arnold, MD (University of Pennsylvania, Site Investigator), Jason H. Karlawish, MD, (University of Pennsylvania, Site Investigator), David Wolk, MD (University of Pennsylvania, Site Investigator), Charles D. Smith, MD (University of Kentucky, Site Investigator), Greg Jicha, MD (University of Kentucky, Site Investigator), Peter Hardy, PhD (University of Kentucky, Site Investigator), Oscar L. Lopez, MD (University of Pittsburgh, Site Investigator), MaryAnn Oakley, MA (University of Pittsburgh, Site Investigator), Donna M. Simpson, CRNP, MPH (University of Pittsburgh, Site Investigator), Anton P. Porsteinsson, MD (University of Rochester Medical Center, Site Investigator), Bonnie S. Goldstein, MS, NP (University of Rochester Medical Center, Site Investigator), Kim Martin, RN (University of Rochester Medical Center, Site Investigator), Kelly M. Makino, BS (University of Rochester Medical Center, Past Site Investigator), M. Saleem Ismail, MD (University of Rochester Medical Center, Past Site Investigator), Connie Brand, RN (University of Rochester Medical Center, Past Site Investigator), Ruth A. Mulnard, DNSc, RN, FAAN (University of California, Irvine, Site Investigator), Gaby Thai, MD (University of California, Irvine, Site Investigator), Catherine Mc-Adams-Ortiz, MSN, RN, A/GNP (University of California, Irvine, Site Investigator), Kyle Womack, MD (University of Texas Southwestern Medical School, Site Investigator), Dana Mathews, MD, PhD (University of Texas Southwestern Medical School, Site Investigator), Mary Quiceno, MD (University of Texas Southwestern Medical School, Site Investigator), Ramon Diaz-Arrastia, MD, PhD (University of Texas Southwestern Medical School, Past Site Investigator), Richard King, MD (University of Texas Southwestern Medical School, Past Site Investigator), Myron Weiner, MD (University of Texas Southwestern Medical School, Past Site Investigator), Kristen Martin-Cook, MA (University of Texas Southwestern Medical School, Past Site Investigator), Michael DeVous, PhD (University of Texas Southwestern Medical School, Past Site Investigator), Allan I. Levey, MD, PhD (Emory University, Site Investigator), James J. Lah, MD, PhD (Emory University, Site Investigator), Janet S. Cellar, DNP, PMHCNS-BC (Emory University, Site Investigator), Jeffrey M. Burns, MD (University of Kansas, Medical Center, Site Investigator), Heather S. Anderson, MD (University of Kansas, Medical Center, Site Investigator), Russell H. Swerdlow, MD (University of Kansas, Medical Center, Site Investigator), Liana Apostolova, MD (University of California, Los Angeles, Site Investigator), Po H. Lu, PsyD (University of California, Los Angeles, Site Investigator), George Bartzokis, MD (University of California, Los Angeles, Past Site Investigator), Daniel H.S. Silverman, MD, PhD (University of California, Los Angeles, Past Site Investigator), Neill R Graff-Radford, MBBCH, FRCP (London) (Mayo Clinic, Jacksonville, Site Investigator), Francine Parfitt, MSH, CCRC (Mayo Clinic, Jacksonville, Site Investigator), Heather Johnson, MLS, CCRP (Mayo Clinic, Jacksonville, Site Investigator), Martin R. Farlow, MD (Indiana University, Site Investigator), Ann Marie Hake, MD, Brandy R. Matthews, MD (Indiana University, Site Investigator), Scott Herring, RN (Indiana University, Past Site Investigator), Christopher H. van Dyck, MD (Yale University School of Medicine, Site Investigator), Richard E. Carson, PhD (Yale University School of Medicine, Site Investigator), Martha G. MacAvoy, PhD (Yale University School of Medicine, Site Investigator), Howard Chertkow, MD (McGill Univ., Montreal-Jewish General Hospital, Site Investigator), Howard Bergman, MD (McGill Univ., Montreal-Jewish General Hospital, Site Investigator), Chris Hosein, MEd (McGill Univ., Montreal-Jewish General Hospital, Site Investigator), Sandra Black, MD, FRCPC (Sunnybrook Health Sciences, Ontario, Site Investigator), Bojana Stefanovic, PhD (Sunnybrook Health Sciences, Ontario, Site Investigator), Curtis Caldwell, PhD (Sunnybrook Health Sciences, Ontario, Site Investigator), Ging-Yuek Robin Hsiung, MD, MHSc, FRCPC (U.B.C. Clinic for AD & Related Disorders, Site Investigator), Howard Feldman, MD, FRCPC (U.B.C. Clinic for AD & Related Disorders, Site Investigator), Benita Mudge, BS (U.B.C. Clinic for AD & Related Disorders, Site Investigator), Michele Assaly, MA (U.B.C. Clinic for AD & Related Disorders, Past Site Investigator), Andrew Kertesz, MD (Cognitive Neurology - St. Joseph’s, Ontario, Site Investigator), John Rogers, MD (Cognitive Neurology - St. Joseph’s, Ontario, Site Investigator), Dick Trost, PhD (Cognitive Neurology - St. Joseph’s, Ontario, Site Investigator), Charles Bernick, MD (Cleveland Clinic Lou Ruvo Center for Brain Health, Site Investigator), Donna Munic, PhD (Cleveland Clinic Lou Ruvo Center for Brain Health, Site Investigator), Diana Kerwin, MD (Northwestern University, Site Investigator), Marek-Marsel Mesulam, MD (Northwestern University, Site Investigator), Kristina Lipowski, BA (Northwestern University, Site Investigator), Chuang-Kuo Wu, MD, PhD (Northwestern University, Past Site Investigator), Nancy Johnson, PhD (Northwestern University, Past Site Investigator), Carl Sadowsky, MD (Premiere Research Inst (Palm Beach Neurology), Site Investigator), Walter Martinez, MD (Premiere Research Inst (Palm Beach Neurology), Site Investigator), Teresa Villena, MD (Premiere Research Inst (Palm Beach Neurology), Site Investigator), Raymond Scott Turner, MD, PhD (Georgetown University Medical Center, Site Investigator), Kathleen Johnson, NP (Georgetown University Medical Center, Site Investigator), Brigid Reynolds, NP (Georgetown University Medical Center, Site Investigator), Reisa A. Sperling, MD (Brigham and Women’s Hospital, Site Investigator), Keith A. Johnson, MD (Brigham and Women’s Hospital, Site Investigator), Gad Marshall, MD (Brigham and Women’s Hospital, Past Site Investigator), Meghan Frey (Brigham and Women’s Hospital, Past Site Investigator), Jerome Yesavage, MD (Stanford University, Site Investigator), Joy L. Taylor, PhD (Stanford University, Site Investigator), Barton Lane, MD (Stanford University, Site Investigator), Allyson Rosen, PhD (Stanford University, Past Site Investigator), Jared Tinklenberg, MD (Stanford University, Past Site Investigator), Marwan Sabbagh, MD, FAAN, CCRI (Banner Sun Health Research Institute, Site Investigator), Christine Belden, PsyD (Banner Sun Health Research Institute, Site Investigator), Sandra Jacobson, MD (Banner Sun Health Research Institute, Site Investigator), Neil Kowall, MD (Boston University, Site Investigator), Ronald Killiany, PhD (Boston University, Site Investigator), Andrew E. Budson, MD (Boston University, Site Investigator), Alexander Norbash, MD (Boston University, Past Site Investigator), Patricia Lynn Johnson, BA (Boston University, Past Site Investigator), Thomas O. Obisesan, MD, MPH (Howard University, Site Investigator), Saba Wolday, MSc (Howard University, Site Investigator), Salome K. Bwayo, PharmD (Howard University, Past Site Investigator), Alan Lerner, MD (Case Western Reserve University, Site Investigator), Leon Hudson, MPH (Case Western Reserve University, Site Investigator), Paula Ogrocki, PhD (Case Western Reserve University, Site Investigator), Evan Fletcher, PhD (University of California, Davis – Sacramento, Site Investigator), Owen Carmichael, PhD (University of California, Davis – Sacramento, Site Investigator), John Olichney, MD (University of California, Davis – Sacramento, Site Investigator), Charles DeCarli, MD (University of California, Davis – Sacramento, Past Site Investigator), Smita Kittur, MD (Neurological Care of CNY, Site Investigator), Michael Borrie, MB ChB (Parkwood Hospital, Site Investigator), T-Y Lee, PhD (Parkwood Hospital, Site Investigator), Dr Rob Bartha, PhD (Parkwood Hospital, Site Investigator), Sterling Johnson, PhD (University of Wisconsin, Site Investigator), Sanjay Asthana, MD (University of Wisconsin, Site Investigator), Cynthia M. Carlsson, MD (University of Wisconsin, Site Investigator), Steven G. Potkin, MD (University of California, Irvine – BIC, Site Investigator), Adrian Preda, MD (University of California, Irvine – BIC, Site Investigator), Dana Nguyen, PhD (University of California, Irvine – BIC, Site Investigator), Pierre Tariot, MD (Banner Alzheimer’s Institute, Site Investigator), Adam Fleisher, MD (Banner Alzheimer’s Institute, Site Investigator), Stephanie Reeder, BA (Banner Alzheimer’s Institute, Site Investigator), Vernice Bates, MD (Dent Neurologic Institute, Site Investigator), Horacio Capote, MD (Dent Neurologic Institute, Site Investigator), Michelle Rainka, PharmD, CCRP (Dent Neurologic Institute, Site Investigator), Douglas W. Scharre, MD (Ohio State University, Site Investigator), Maria Kataki, MD, PhD (Ohio State University, Site Investigator), Earl A. Zimmerman, MD (Albany Medical College, Site Investigator), Dzintra Celmins, MD (Albany Medical College, Site Investigator), Alice D. Brown, FNP (Albany Medical College, Past Site Investigator), Godfrey D. Pearlson, MD (Hartford Hosp, Olin Neuropsychiatry Research Center, Site Investigator), Karen Blank, MD (Hartford Hosp, Olin Neuropsychiatry Research Center, Site Investigator), Karen Anderson, RN (Hartford Hosp, Olin Neuropsychiatry Research Center, Site Investigator), Andrew J. Saykin, PsyD (Dartmouth-Hitchcock Medical Center, Site Investigator), Robert B. Santulli, MD (Dartmouth-Hitchcock Medical Center, Site Investigator), Eben S. Schwartz, PhD (Dartmouth-Hitchcock Medical Center, Site Investigator), Kaycee M. Sink, MD, MAS (Wake Forest University Health Sciences, Site Investigator), Jeff D. Williamson, MD, MHS (Wake Forest University Health Sciences, Site Investigator), Pradeep Garg, PhD (Wake Forest University Health Sciences, Site Investigator), Franklin Watkins, MD (Wake Forest University Health Sciences, Past Site Investigator), Brian R. Ott, MD (Rhode Island Hospital, Site Investigator), Henry Querfurth, MD (Rhode Island Hospital, Site Investigator), Geoffrey Tremont, PhD (Rhode Island Hospital, Site Investigator), Stephen Salloway, MD, MS (Butler Hospital, Site Investigator), Paul Malloy, PhD (Butler Hospital, Site Investigator), Stephen Correia, PhD (Butler Hospital, Site Investigator), Howard J. Rosen, MD (UC San Francisco, Site Investigator), Bruce L. Miller, MD (UC San Francisco, Site Investigator), Jacobo Mintzer, MD, MBA (Medical University South Carolina, Site Investigator), Crystal Flynn Longmire, PhD (Medical University South Carolina, Site Investigator), Kenneth Spicer, MD, PhD (Medical University South Carolina, Site Investigator), Elizabether Finger, MD (St. Joseph’s Health Care, Site Investigator), Irina Rachinsky, MD (St. Joseph’s Health Care, Site Investigator), John Rogers, MD (St. Joseph’s Health Care, Site Investigator), Andrew Kertesz, MD (St. Joseph’s Health Care, Past Site Investigator), Dick Drost, MD (St. Joseph’s Health Care, Past Site Investigator), Nunzio Pomara, MD (Nathan Kline Institute, Site Investigator), Raymundo Hernando, MD (Nathan Kline Institute, Site Investigator), Antero Sarrael, MD (Nathan Kline Institute, Site Investigator), Susan K. Schultz, MD (University of Iowa, Site Investigator), Laura L. Boles Ponto, PhD (University of Iowa, Site Investigator), Hyungsub Shim, MD (University of Iowa, Site Investigator), Karen Elizabeth Smith, RN (University of Iowa, Site Investigator), Norman Relkin, MD, PhD (Cornell University), Gloria Chaing, MD (Cornell University), Lisa Raudin, PhD (Cornell University), Amanda Smith, MD (University of South Florida), Kristin Fargher, MD (University of South Florida), Balebail Ashok Raj, MD (University of South Florida).

## Notes

### Competing Interest Statement

The authors have declared no competing interest.

### Author Declarations

All data are available on the ADNI websites upon demand (http://adni.loni.usc.edu/data-samples/access-data/).

